# Circulation of DENV-2 serotype associated with increased risk of cumulative incidence of severe dengue and dengue with warning signs: A 16-year retrospective study in Peru

**DOI:** 10.1101/2024.05.02.24306735

**Authors:** Jorge L. Cañari-Casaño, Valerie A. Paz-Soldan, Andres G. Lescano, Amy C. Morrison

**Affiliations:** One Health Research Unit. School of Public Health and Administration, Universidad Peruana Cayetano Heredia, Lima, Peru; Tropical Medicine and Infectious Disease, Tulane School of Public Health and Tropical Medicine, New Orleans, Louisiana, US; Emerge, Emerging Diseases and Climate Change Research Unit, School of Public Health and Administration, Universidad Peruana Cayetano Heredia, Lima, Peru; Department of Pathology, Microbiology, and Immunology, School of Veterinary Medicine, University of California, Davis, California, US

**Keywords:** Dengue, Serotypes, dengue severity

## Abstract

**Background:** Dengue poses a significant public health challenge in Peru and other endemic countries worldwide. While severe dengue is known to be associated with secondary infection at the individual level, the factors that elevate the risk of severe dengue at the population level remain poorly understood. This study leverages over 16 years of secondary data from a Peruvian dengue surveillance system to assess which type of serotype-specific circulation is associated with an increased risk of cumulative incidence of severe dengue or dengue with warning signs (SD-DWS).

**Methodology:** This is a retrospective analysis of secondary data using the Peruvian Ministry of Health databases of dengue cases and serotyping. A mixed negative binomial regression model for repeated measures over time was employed to estimate the association between the cumulative incidence of reported SD-DWS cases per 100,000 inhabitants and serotype-specific circulation. Crude and adjusted incidence ratios (IRR) were estimated.

**Principal findings:** The study analyzed data from 2007 to 2022 across 19 regions of Peru, totaling 304 region-years. Data from nearly 58,000 serotype identification reports and 57,966 cases of SD-DWS were analyzed. The regions with most cumulative incidence of SD-DWS per 100,000 inhabitants during 2007 to 2022 were Madre de Dios (3859), Loreto (1518), Ucayali (1492), Tumbes (1335), and Piura (722). The adjusted model revealed a higher risk of cumulative incidence of SD-DWS when there was specific circulation of DENV-123 (aIRR 7.57 CI 4.00 – 14.31), DENV-12 (aIRR 4.66 CI 2.57 – 8.44), DENV-23 (aIRR 3.55 CI 1.75 – 7.21), or when there was circulation of DENV-2 alone or co-circulating with other serotypes (aIRR 27.7 CI 15.46 -49.63).

**Conclusions:** Circulation of DENV-2 was associated with higher average incidence rate ratios of SD-DWS.

**Author summary:** We investigated how the circulation of different dengue virus (DENV) serotypes are associated with the incidence of severe dengue and dengue with warning signs in Peru, a country where dengue is endemic. We analyzed 16 years of data from the dengue surveillance system, including nearly 58,000 serotype identification reports and 57,966 cases of severe dengue and dengue with warning signs. We found that regions with specific circulation of DENV-2, either alone or in combination with other serotypes, had higher incidence rates of severe dengue and dengue with warning signs. Our findings highlight the importance of monitoring DENV serotype circulation to manage and prevent severe dengue, especially in regions where DENV-2 is prevalent.

## Introduction

Dengue, a significant public health issue, is endemic in Peru, as in other tropical climates. It is caused by an RNA virus with four known serotypes (DENV-1, DENV-2, DENV-3, and DENV-4) [1]. The dengue virus (DENV) is primarily transmitted through the bite of an infected female mosquito of the *Aedes* genus: in Peru, it is mainly transmitted by *Aedes aegypti* [2,3].

Most of the dengue research in Peru has been carried out in the Amazonian city of Iquitos. These studies have shown that since the reintroduction of dengue in Peru in 1990, all four DENV serotypes (including the introduction of new genotypes) have circulated [4–6]. In addition, Peru has experienced several epidemic outbreaks (2001/2002, 2011/2012, 2017, and 2023) with a high number of cases of severe dengue (SD) or dengue with warning signs (DWS) (SD-DWS), sometimes overwhelming or collapsing health systems [2,7].

For an individual, a secondary DENV infection with a heterologous serotype is one of the primary risk factors for severe dengue [8–12]. At the population level, the risk of severe disease may be associated with 1) an increase in the population susceptible to second infections, 2) the sequential or co-circulation of multiple DENV serotypes [13], and 3) a rise in the overall force of infection, increasing the number of all cases across the clinical spectrum of disease and the probability that individuals susceptible to a second infection become infected. Additionally, inadequate disease surveillance and vector control, inadequate or improper case management, all contribute to the public health consequences of dengue outbreaks, and particularly, of severe disease.

Due to the low prevalence of severe dengue (less than 1% of total dengue cases [3]), identification of specific risk factors for SD-DWS requires multiple years (decades) of surveillance to achieve adequate statistical power for analyses. Our study leverages 16 years of Peruvian dengue surveillance system data to evaluate potential serotype-related risk factors (what and how many serotypes are circulating) associated with increased incidence of SD-DWS.

## Methods

### Ethics statement

Our secondary data protocol was approved by the institutional review board of the Peruvian University Cayetano Heredia (SIDISI 207993, approval record number 079-01-22). The anonymized data was obtained through the public information system of the Peruvian Ministry of Health.

### Data sources

Data was acquired from the National Center for Epidemiology of Peru (CDC-Peru), a branch of the Ministry of Health. We extracted all reported dengue cases and serotyping data collected between 2007 and 2022. Reporting of dengue cases is mandatory for both public and private health centers nationwide [14]. This surveillance system reports both confirmed and probable cases. Confirmed cases are those with laboratory confirmation by DENV isolation, RT-PCR, NS1 antigen, detection of IgM antibodies for dengue in a single sample, and/or evidence of IgM seroconversion in paired samples. Probable cases correspond to those that occur during outbreaks, meet case definitions set by the World Health Organization [15] or by epidemiological link for patients who had direct contact with confirmed cases [14]. Overall, 90% of cases in our database were classified as confirmed and 10% represented probable cases.

A small proportion of these confirmed cases are analyzed in regional reference laboratories to identify the circulating serotype using multiplex RT-PCR [16]. Subsequently, the serotype identification reports are registered in the database system of the Peruvian Public Health Laboratory Network (NETLAB) [17] of the Ministry of Health. Dengue case data and serotype identification data were requested from the Ministry of Health through their public access website [18].

### Study variables

Our principal outcome variable was the cumulative incidence of SD-DWS per 100,000 inhabitants, associated with explanatory serotype-specific circulation variables. We calculated the cumulative incidence of SD-DWS per 100,000 inhabitants for each region and year by dividing the total number of SD-DWS by the projected population, estimated by year by the National Institute of Statistics and Informatics (INEI) using census data. We also estimated a global summary of the cumulative incidence of SD-DWS per 100,000 inhabitants for each region over the 16-year study period, by dividing – per region (Peruvian state) – the total number of SD-DWS cases by the average annual projected population. Serotype-specific circulation variables were created based on the total possible combinations of each serotype detected per region, or their combination (DENV-1, DENV-2, DENV-3, DENV-4, DENV-12, DENV-13, DENV-14, DENV-23, DENV-24, DENV-34, DENV-123, DENV-124, DENV-134, DENV-234, and DENV-1234). Each of these variables were coded as 0, if there was no presence of circulation, or 1, if there was specific presence of such circulation. Additionally, to assess which serotype type has the highest risk for elevated cumulative DS-DWS incidence, we created 4 additional variables (circulation of DENV-1 or its combination, circulation of DENV-2 or its combination, circulation of DENV-3 or its combination, and circulation of DENV-4 or its combination).

Additionally, for each region, we adjusted for poverty level and total number of primary care health facilities per population. The poverty level indicator was based on the INEI categorization of “poverty with at least one unmet basic need”. A family is “poor” if it has at least 1 of the following characteristics: 1) home with inadequate construction (exterior walls made of straw mats or “quincha”, or stone with mud or wood, and dirt floor), 2) overcrowded home (more than 3 to 4 people per room), 3) home without indoor plumbing, 4) home with children who do not attend school, and 5) home with high economic dependence [19].

Regarding number of health facilities, in Peru there are 3 levels of health facilities based on the complexity of the care and services: 1) primary care facilities, 2) general hospitals or clinics, 3) specialized hospitals and institutes (e.g., national oncological institute) [20]. We extracted the total number of primary care facilities per region from the Ministry of Health databases [18]. During outbreaks, primary care facilities provide care to patients suspected of dengue, provide basic care, and prevent progression to severe forms of the disease. Finally, per region, the total number of primary health facilities was divided by the projected population.

### Population and inclusion/exclusion criteria

The study population consisted of Peruvian regions (states) with reports of SD and/or DWS and dengue serotype identification from 2007 to 2022. Regions without reports of SD-DWS cases and without identification of circulating serotypes were excluded from the analysis.

### Analytical Approach

For the descriptive analysis, we created summary tables and trend graphs of our variables of interest. For the main analysis, we explored the association between the cumulative incidence of SD-DWS per 100,000 inhabitants with each of 16 possible combinations of serotype circulation (see study variable description above). In a second approach, we explored the association of SD-DWS per 1000,000 inhabitants by the 4 variables that indicated the circulation of each serotype or its combination. For both analyses, we considered confounding adjustment variables (defined a priori by epidemiological criteria), such as the poverty level variable (at the region-year level) and the number of primary health centers per population per region. Given that our outcome variable had overdispersion, we used mixed negative binomial regression models for repeated measures over time [21] to estimate crude and adjusted cumulative incidence ratios (IRR) using the “menbreg” command in Stata (versión 17.0, StataCorp LLC, College Station, TX), for this we also evaluated multicollinearity among the predictors. The analyses will be performed with a significance level (α) of 0.05 and a confidence level of 95%. The statistical programs used were Stata and R software (version 4.3.2.).

## Results

Overall, 19 regions met our inclusion criteria for the years 2007 to 2022, for a total of 304 region-years included in our analysis. Five regions (Apurímac, Arequipa, Huancavelica, Moquegua, and Tacna) were excluded from the analysis because they either had no reports of SD-DWS cases or no serotype information was available.

### Dengue cases and cumulative incidence of SD-DWS

Five regions account for 65% of reported dengue cases, and almost 80% of both DWS and SD in Peru (Table 1). Based on WHO definitions, 87.1%, 12.5%, and only 0.4% were classified as dengue without warning signs, DWS, and SD, respectively (Table 1). Some of the regions with highest cumulative dengue incidence were different than those with highest absolute numbers, with four having greater than 1,334 cases of SD-DWS per 100,000 inhabitants (Table 2 and Fig 1).

**Table 1.** Reported dengue cases by WHO classification by region from 2007 to 2022.

| Regions | Dengue without warning signs | Dengue with warning signs | Dengue severe | Total cases of dengue | Percent by region |
| --- | --- | --- | --- | --- | --- |
| Piura | 98,293 | 13,266 | 330 | 111,889 | 24.9% |
| Loreto | 67,712 | 14,188 | 517 | 82,417 | 18.3% |
| Ucayali | 30,910 | 7,442 | 305 | 38,657 | 8.6% |
| Madre de Dios | 25,912 | 5,342 | 257 | 31,511 | 7.0% |
| San Martín | 21,651 | 4,218 | 146 | 26,015 | 5.8% |
| Ica | 21,167 | 601 | 33 | 21,801 | 4.9% |
| Tumbes | 20,751 | 3,015 | 31 | 23,797 | 5.3% |
| Junín | 16,295 | 2,039 | 65 | 18,399 | 4.1% |
| La Libertad | 15,653 | 950 | 36 | 16,639 | 3.7% |
| Cajamarca | 13,494 | 1,870 | 36 | 15,400 | 3.4% |
| Lambayeque | 13,148 | 231 | 28 | 13,407 | 3.0% |
| Cusco | 10,177 | 271 | 22 | 10,470 | 2.3% |
| Amazonas | 9,562 | 509 | 38 | 10,109 | 2.3% |
| Ayacucho | 8,603 | 270 | 20 | 8,893 | 2.0% |
| Huánuco | 7,087 | 1,042 | 33 | 8,162 | 1.8% |
| Ancash | 6,069 | 300 | 20 | 6,389 | 1.4% |
| Lima | 3,544 | 265 | 5 | 3,814 | 0.9% |
| Pasco | 1,714 | 205 | 17 | 1,936 | 0.4% |
| Puno | 175 | 3 | 0 | 178 | 0.04% |
| Total | 391,917 | 56,027 | 1,939 | 449,883 | 100% |
| N (%) | (87.1%) | (12.5%) | (0.4%) |  |  |

**Table 2.** Cumulative incidence of severe dengue and dengue with warning signs (SD-DWS) per 100,000 inhabitants from 2007 to 2022.

| Regions | Average population | SD-DWS cases | Cumulative incidence per 100,000 inhabitants |
| --- | --- | --- | --- |
| Madre de Dios | 145,078 | 5,599 | 3,859 |
| Loreto | 968,879 | 14,705 | 1,518 |
| Ucayali | 519,088 | 7,747 | 1,492 |
| Tumbes | 228,246 | 3,046 | 1,335 |
| Piura | 188,2302 | 13,596 | 722 |
| San Martín | 825,404 | 4,364 | 529 |
| Junín | 130,9566 | 2,104 | 161 |
| Huánuco | 763,715 | 1,075 | 141 |
| Cajamarca | 143,2905 | 1,906 | 133 |
| Amazonas | 413,967 | 547 | 132 |
| Pasco | 275,340 | 222 | 81 |
| Ica | 855,401 | 634 | 74 |
| La Libertad | 1,838,790 | 986 | 53 |
| Ayacucho | 652,514 | 290 | 44 |
| Ancash | 1,129,717 | 320 | 28 |
| Cusco | 1,278,655 | 293 | 23 |
| Lambayeque | 1,221,466 | 259 | 21 |
| Lima | 9,603,276 | 270 | 3 |
| Puno | 1,253,074 | 3 | <1 |
| **Total Average** | **26,597,384** | **57,966** | **218** |

**Figure 1.**
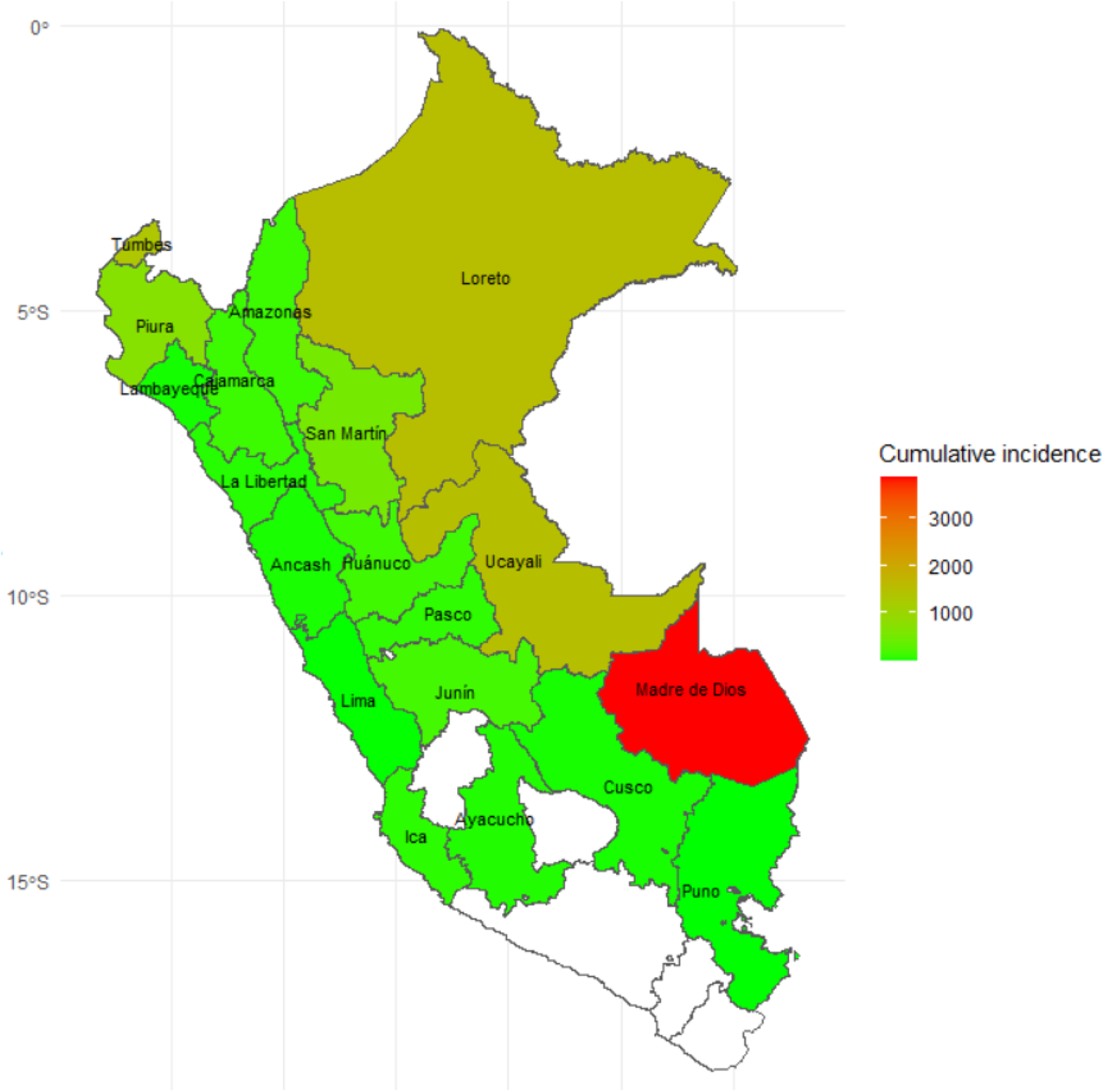
Cumulative incidence of SD-DWS per 100,000 inhabitants by region from 2007 to 2022

### Serotype circulation

From 2007 to 2022, 58,748 samples were serotyped (14.5% of laboratory-confirmed dengue cases) and registered in the NETLAB-INS system. During the observation period, DENV-2 serotype (52.8%) was the most frequently observed, followed by DENV-1 (37.4%), and to a lesser extent DENV-3 (6.9%) and DENV-4 serotypes (2.9%) (Table 3). In most regions, we observed consistent circulation of one or more serotypes over time, with one serotype predominating over others; additionally, after novel serotype introductions or reintroductions of a serotype that had not circulated at high rates for a few years, we could observe displacement of one serotype by another. For example, in Loreto in 2007, DENV-3 was the predominant circulating serotype; it was displaced by DENV-4 in 2008 until 2010, and then after a brief period when DENV-1, DENV-2, and DENV-4 were co-circulating at the same time, DENV-2 caused a dramatic outbreak and became the predominant serotype in the region for nearly a decade (S1 Fig).

**Table 3.**
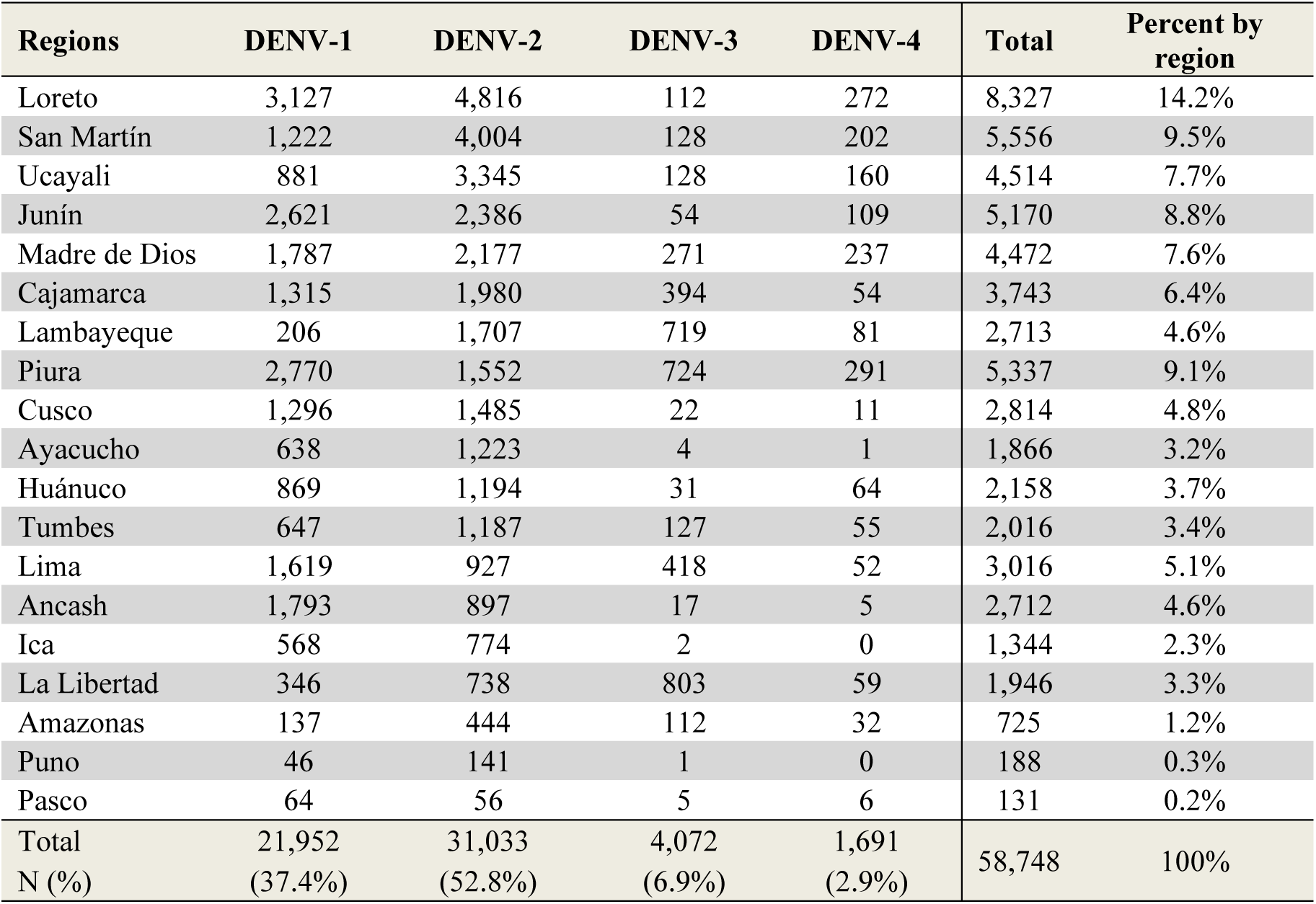
Circulating dengue serotypes by region and year registered in the NETLAB-INS system from 2007 to 2022.

When examining different combinations of serotypes, of the 304 region-years analyzed, DENV-12 was the most commonly observed combination of circulating serotypes, which circulated in 74 region-years (24.3%), followed by DENV-2 (35 region-years), DENV-23 (29 region-years), DENV-123 (27 region-years), and DENV-134 (26 region-years), while in 37 region-years (12.2%) no serotype circulation was reported (S1 Table).

### Serotype circulation attributed to severe dengue

Focusing on severe dengue cases alone, the highest frequency of reported SD cases occurred when there was circulation of DENV-12 (37%), followed by DENV-123 (19.7%) and DENV-23 (12.6%), noting that more SD cases were observed with the involvement of DENV-2 in some combination or alone (Fig 2).

**Figure 2.**
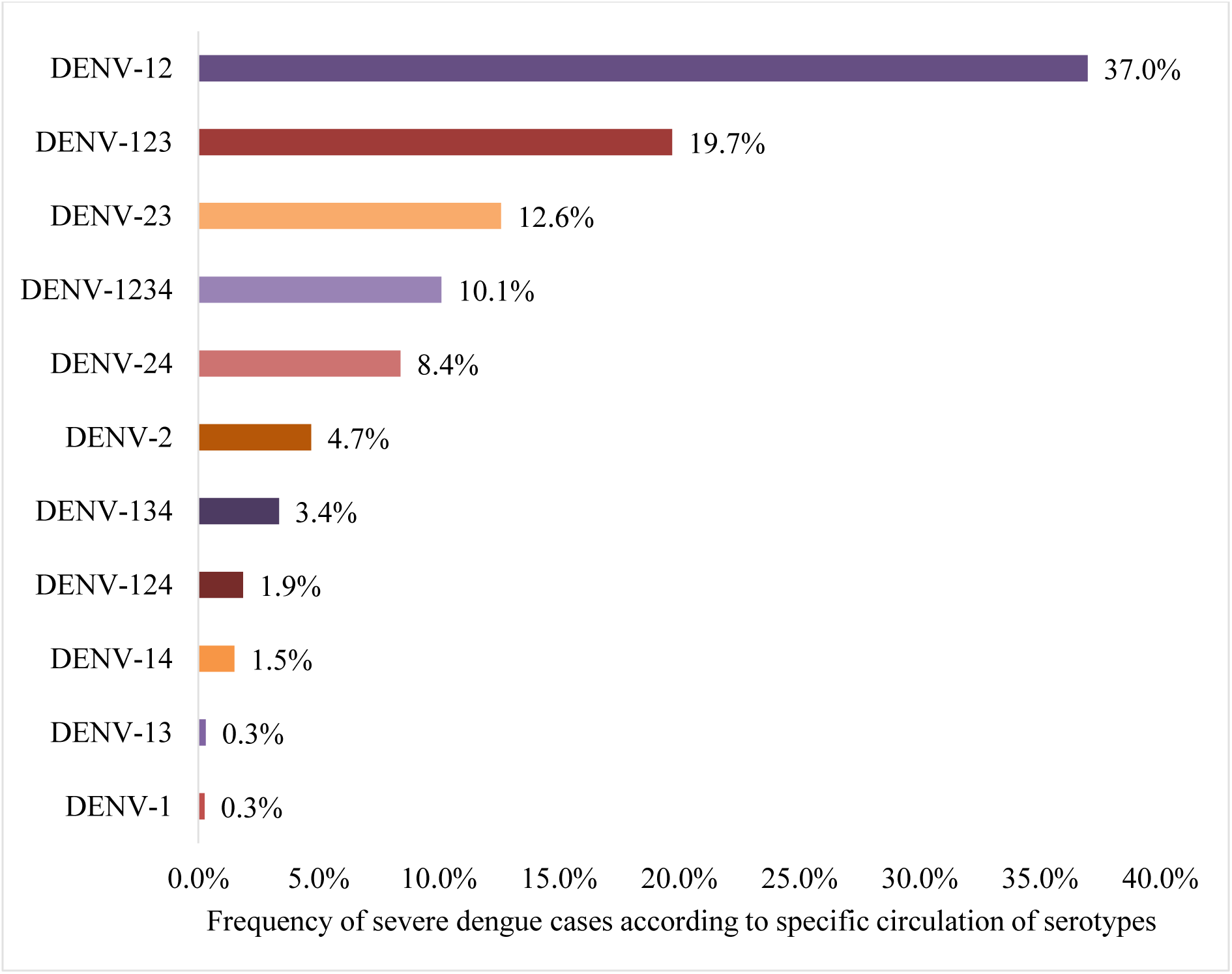
Frequency of severe dengue cases attributed to specific serotype circulation.

### Association between SD-DWS and serotype circulation

Table 4 presents results from our mixed negative binomial regression models for the association between specific serotype circulation and the cumulative incidence of SD-DWS. In the adjusted model, we observed that there is a higher risk for increased cumulative incidence of SD-DWS in regions where DENV-1, DENV-2, and DENV-3 cocirculated (DENV-123); the average incidence ratio per 100,000 inhabitants increases by 7.6 times. Note that the circulating types of serotypes DENV-124, DENV-13, DENV-24, DENV-4, DENV-3, DENV-34, DENV-234, DENV-14, and DENV-1234, were not included in the adjusted model because they circulated infrequently (circulated in less than 6% of the study period, S1 Table) and because their inclusion yielded wide confidence intervals. However, in the adjusted analysis, when all types of serotype circulation were included, the circulation of serotype 2 or its combination showed a high-risk factor for the incidence of SD-DWS (S2 Table). Hence, we conducted a separate analysis to evaluate the cumulative incidence risk of SD-DWS due to the circulation of DENV-2 alone or in combination with other serotypes and found the average incidence ratio per 100,000 inhabitants increases up to 27.7 times (Table 5).

**Table 4.** Association between specific serotype circulation and cumulative incidence of severe dengue and/or dengue with warning signs.

| Variables | Cumulative Incidence of SD-DWS |  |  |  |  |  |  |  |  |  |
| --- | --- | --- | --- | --- | --- | --- | --- | --- | --- | --- |
|  | Crude bivariate analysis |  |  |  |  | Multivariate adjusted analysis |  |  |  |  |
|  | IRRc | IC 95% |  |  | p | IRRa | IC 95% |  |  | p |
| Circulation of DENV-123 |  |  |  |  |  |  |  |  |  |  |
| No | Ref. |  |  |  |  | Ref. |  |  |  |  |
| Yes | 3.17 | 1.55 | - | 6.50 | 0.002 | 7.57 | 4.00 | - | 14.31 | <0.001 |
| Circulation of DENV-12 |  |  |  |  |  |  |  |  |  |  |
| No | Ref. |  |  |  |  | Ref. |  |  |  |  |
| Yes | 3.27 | 2.04 | - | 5.25 | <0.001 | 4.66 | 2.57 | - | 8.44 | <0.001 |
| Circulation of DENV-23 |  |  |  |  |  |  |  |  |  |  |
| No | Ref. |  |  |  |  | Ref. |  |  |  |  |
| Yes | 1.22 | 0.58 | - | 2.57 | 0.599 | 3.55 | 1.75 | - | 7.21 | <0.001 |
| Circulation of DENV-2 |  |  |  |  |  |  |  |  |  |  |
| No | Ref. |  |  |  |  | Ref. |  |  |  |  |
| Yes | 0.67 | 0.33 | - | 1.39 | 0.288 | 1.86 | 0.91 | - | 3.83 | 0.091 |
| Circulation of DENV-1 |  |  |  |  |  |  |  |  |  |  |
| No | Ref. |  |  |  |  | Ref. |  |  |  |  |
| Yes | 0.07 | 0.02 | - | 0.21 | <0.001 | 0.22 | 0.07 | - | 0.66 | 0.007 |
| Circulation of DENV-134 |  |  |  |  |  |  |  |  |  |  |
| No | Ref. |  |  |  |  | Ref. |  |  |  |  |
| Yes | 0.01 | 0.01 | - | 0.03 | <0.001 | 0.09 | 0.03 | - | 0.24 | <0.001 |
| Poverty level at the regional level | 0.85 | 0.81 | - | 0.89 | <0.001 | 0.93 | 0.89 | - | 0.97 | 0.001 |
| Total primary health centers per total population per region | 0.97 | 0.93 | - | 1.02 | 0.220 | 1.00 | 0.95 | - | 1.04 | 0.867 |

IRRc: Crude cumulative incidence ratios per 100,000 population. IRRa: Adjusted cumulative incidence ratios per 100,000 population. For repeated measures over time, a mixed negative binomial regression model was used.

**Table 5:**
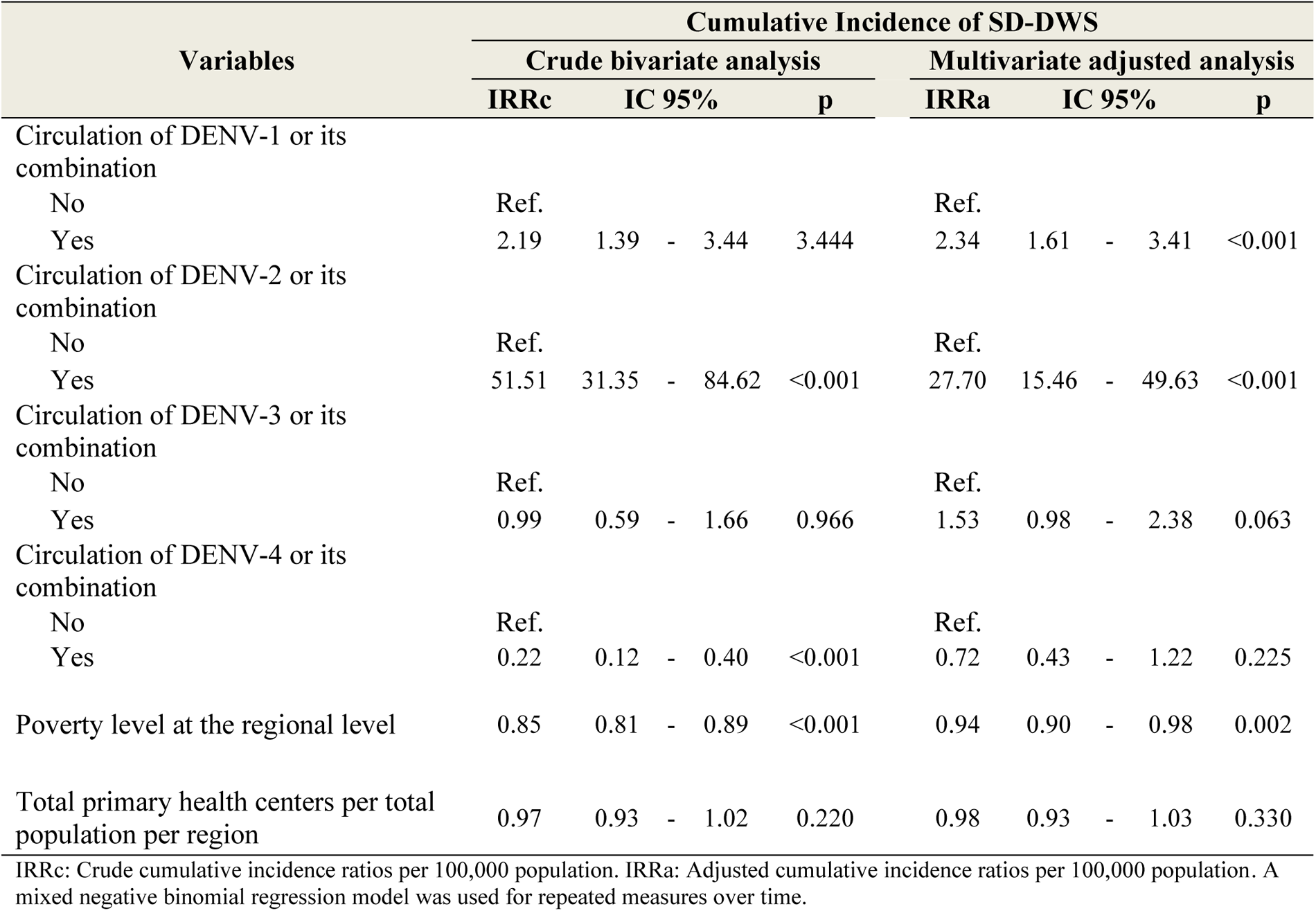
Association between the circulation of each serotype or its combination with the cumulative incidence of severe dengue and/or dengue with warning signs.

### Association between SD-DWS and poverty

We also observed that as regional poverty increased, the cumulative incidence of SD-DWS reduced between 6% (Table 5) and 7% (Table 4). Specifically, in regions with a high level of poverty, such as Loreto and Ucayali, there is a trend of lower cumulative incidence of DS-DWS, while in regions with a low level of poverty, there is a trend of increasing incidence of DS-DWS (S2 Fig.). To explore these results further, we categorized the poverty-level variables into tertiles and re-ran the main model. In this new analysis, we found that regions in the highest tertile (wealthier region) had up to 14.2 times more DS-DWS incidence than regions in the lowest tertile (Table S3).

## Discussion

Our results suggest that the population rates of SD-DWS increase when DENV-2 is one of multiple co-circulating dengue virus serotypes in a region. These findings are consistent with previous studies in other countries that have evidenced a higher risk of severe dengue related to the circulation of the DENV-2 serotype [22–24].

Although our results indicate a higher risk of disease severity due to the presence of DENV-2, we could not determine if this was due to the introduction of a novel genotype (Asian/American) and Cosmopolitan genotypes) because genotyping is limited. Other studies from Peru, however, have described increased disease severity associated with presentations of the 2010 introduction of Asian/American DENV-2 to Iquitos [7,25–27]. The 2019 dengue outbreak in Madre de Dios coincided with the entry of the Cosmopolitan genotype of DENV-2 as well [28]. In 2023, an unprecedented outbreak in both size and areas affected had a particularly devastating impact on the northern coast of Peru, with historic highs of reported SD cases and mortality; once again, it was the circulation of the Cosmopolitan genotype of DENV-2 [2].

The lower cumulative incidence of SD-DWS observed in the context of DENV-1 circulation and the combination of DENV-134 provides additional support for the finding that serotype DENV-2, or its combination is a major risk factor for severe disease. It is important to note, however, that this observation is not equivalent to the conclusion that these other serotypes are protective against severe disease.

We showed that when regional poverty increased, the cumulative incidence of SD-DWS reduced by about 7%. Some studies have found that poor nutritional status can be protective against severe disease at the individual level [29,30]. Many host and environmental factors associated with poverty have multiple and complex impacts on dengue virus transmission, requiring detailed and long-term longitudinal research studies.

This study highlights the importance of diagnostic surveillance of circulating DENV serotypes. As DENV expands to new areas of Peru, as has occurred recently, health personnel with no or limited experience with dengue must be trained on its management and treatment. Shifts in DENV serotypes or genotypes increase the risk of epidemic transmission because the population likely has lower levels of immunity toward a serotype that has been absent for multiple years. Additionally, the increased risk for severe diseases associated with DENV-2 is well established in cohort studies of individuals [22–24], and our study validates this at the population level.

There are various limitations to this study. We used public data aggregated at the population level and lacked detailed information on the identification of the circulating serotype (or its phenotypic variants) for each dengue case reported by the epidemiological surveillance system or the identification of the type of infection (first, second, third, or fourth) of each reported dengue case. The information obtained about the diagnosed dengue cases comes from the CDC-Peru’s epidemiological surveillance system; reported by public and private health systems at all levels of care and based on the diagnostic criteria established by the WHO in 2009 [15]. As a result, variability in the diagnosis/reporting of dengue cases by health personnel is very likely; however, this would result in a non-differential bias, as it would affect all dengue diagnostic groups and all study regions similarly [31]. Additionally, the Peruvian dengue epidemiological surveillance is based on a passive surveillance system: this likely results in underreporting of dengue cases and identification of the circulating serotype, either because most dengue cases are asymptomatic (of every 5 people with dengue, only one person is symptomatic [32]) or due to lack of access to care in the health system (especially in the context of epidemic outbreaks). Finally, an important limitation is that population-level variables that could influence the disease severity have not been taken into account in this study, such as the population at risk of second infections (risk factor for severe dengue at the individual level) or vector control actions carried out regionally (street and home fumigation, water treatment, and breeding site collection campaigns), which are known to help reduce transmission intensity during an epidemic outbreak.

## Acknowledgments

We thank the National Institute of Health of the Peruvian Ministry of Health and the CDC-Peru for sharing data from their epidemiological surveillance system for dengue.

## Data Availability

The final dataset used is available on https://doi.org/10.6084/m9.figshare.25336015.v4

## Supporting information

**S1 Fig.**
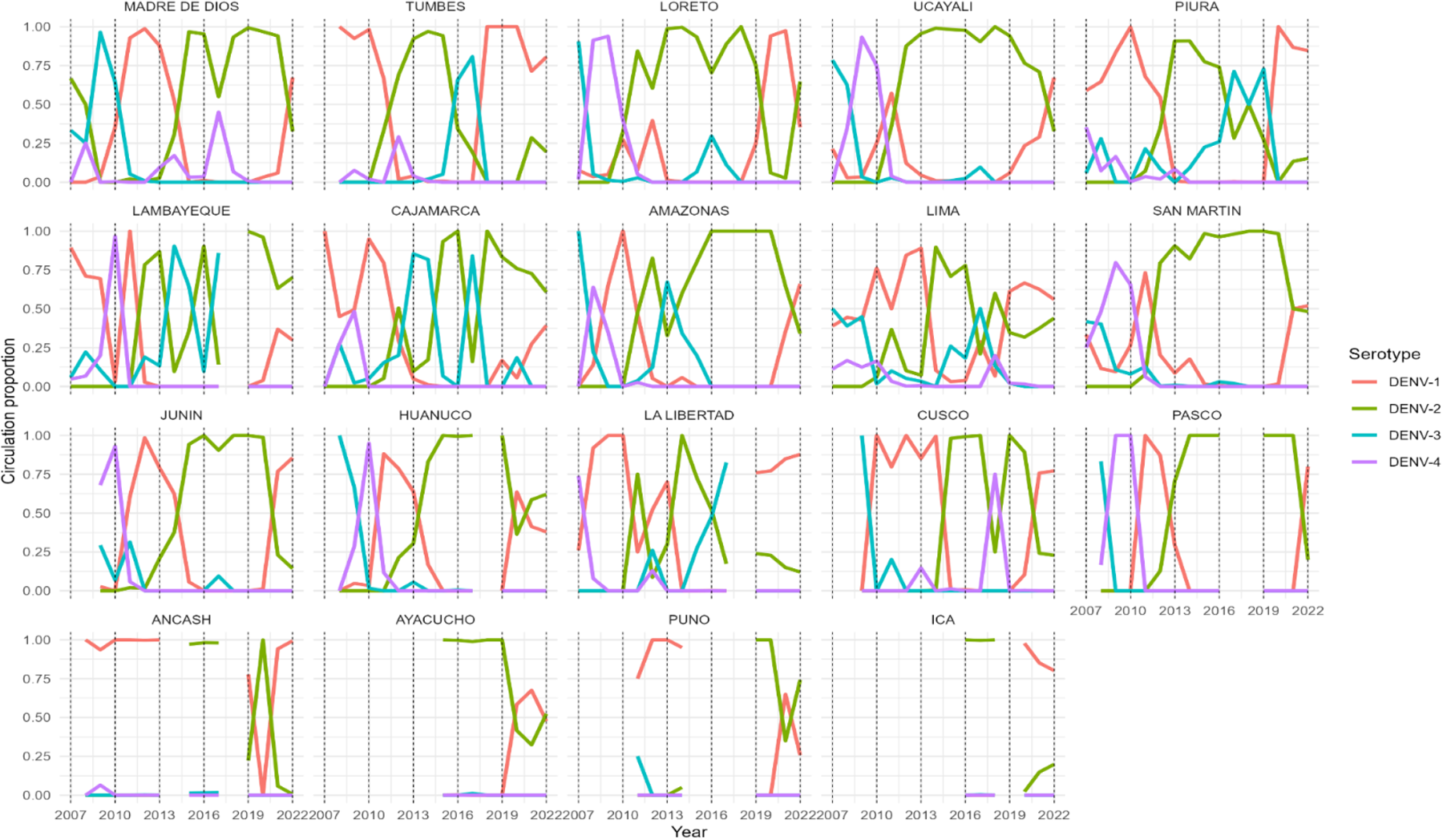
Circulation proportion of dengue serotypes in Peru from 2007 to 2022.

**S2 Fig.**
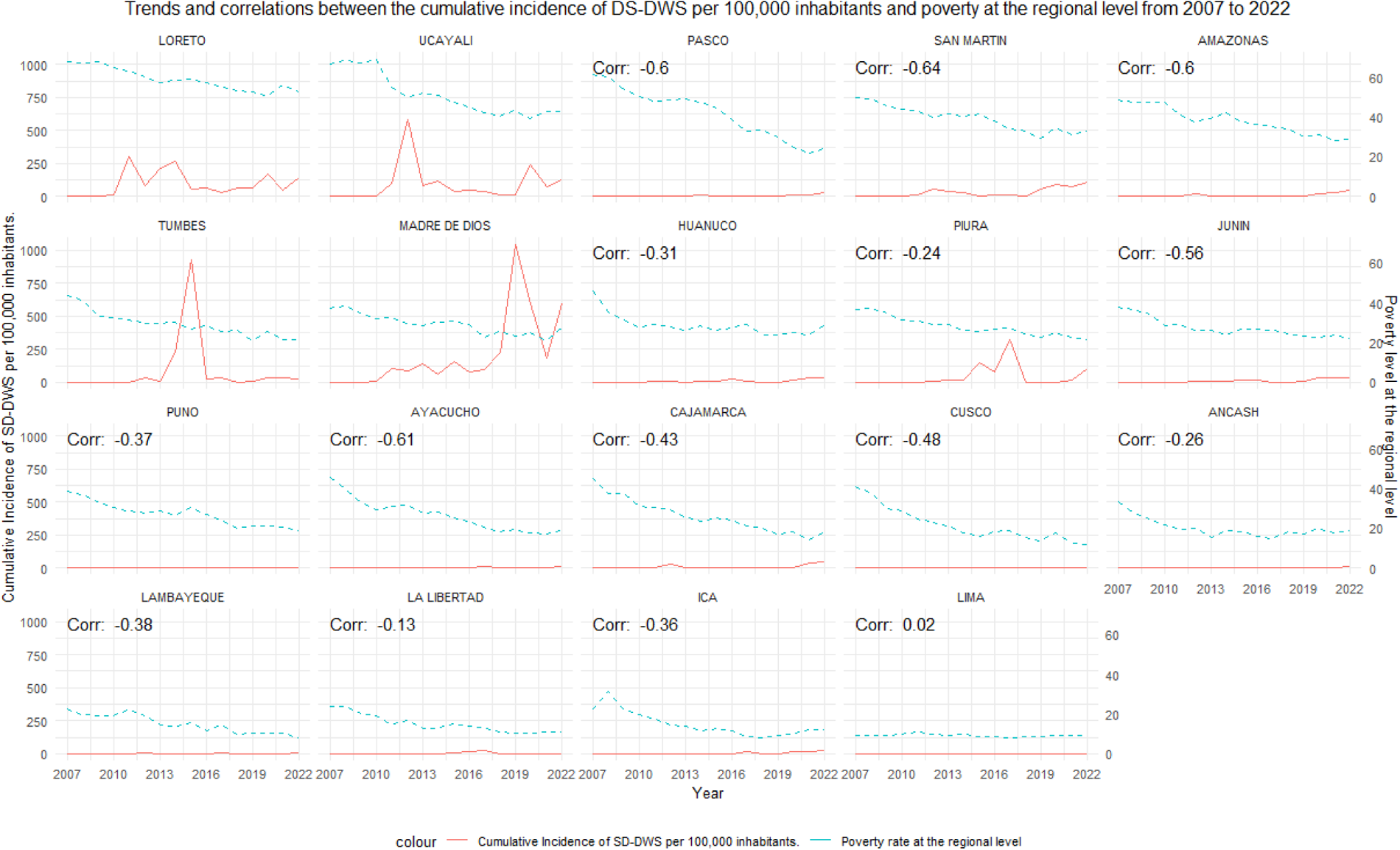
Trends and correlations between the cumulative incidence of SD-DWS per 100,000 inhabitants and poverty at the regional level from 2007 to 2022.

**S1 Table.**
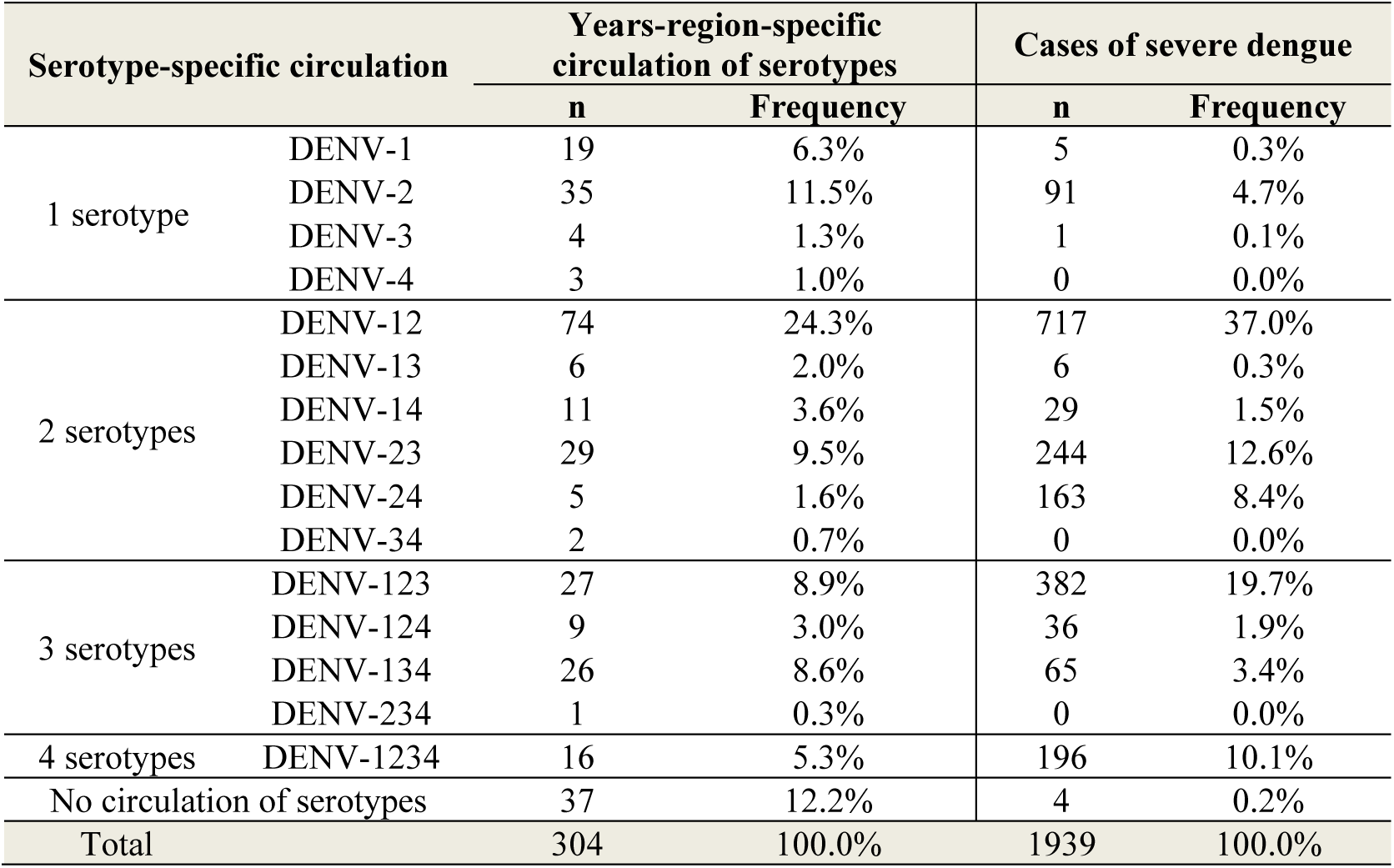
Serotype-specific circulation and severe dengue cases.

| Serotype-specific circulation |  | Years-region-specific circulation of serotypes |  | Cases of severe dengue |  |
| --- | --- | --- | --- | --- | --- |
|  |  | n | Frequency | n | Frequency |
| 1 serotype | DENV-1 | 19 | 6.3% | 5 | 0.3% |
|  | DENV-2 | 35 | 11.5% | 91 | 4.7% |
|  | DENV-3 | 4 | 1.3% | 1 | 0.1% |
|  | DENV-4 | 3 | 1.0% | 0 | 0.0% |
| 2 serotypes | DENV-12 | 74 | 24.3% | 717 | 37.0% |
|  | DENV-13 | 6 | 2.0% | 6 | 0.3% |
|  | DENV-14 | 11 | 3.6% | 29 | 1.5% |
|  | DENV-23 | 29 | 9.5% | 244 | 12.6% |
|  | DENV-24 | 5 | 1.6% | 163 | 8.4% |
|  | DENV-34 | 2 | 0.7% | 0 | 0.0% |
| 3 serotypes | DENV-123 | 27 | 8.9% | 382 | 19.7% |
|  | DENV-124 | 9 | 3.0% | 36 | 1.9% |
|  | DENV-134 | 26 | 8.6% | 65 | 3.4% |
|  | DENV-234 | 1 | 0.3% | 0 | 0.0% |
| 4 serotypes | DENV-1234 | 16 | 5.3% | 196 | 10.1% |
| No circulation of serotypes |  | 37 | 12.2% | 4 | 0.2% |
| Total |  | 304 | 100.0% | 1939 | 100.0% |

**S2 Table.** Association between all serotype-specific circulation types and cumulative incidence of SD-DWS.

| Variables | Cumulative Incidence of SD-DWS |  |  |  |  |
| --- | --- | --- | --- | --- | --- |
|  | Multivariate adjusted analysis |  |  |  |  |
|  | IRRa | IC 95% |  |  | p |
| Circulation of DENV-123 | 243.67 | 53.12 | - | 1117.71 | <0.001 |
| Circulation of DENV-24 | 142.59 | 21.67 | - | 938.21 | <0.001 |
| Circulation of DENV-12 | 128.99 | 29.07 | - | 572.31 | <0.001 |
| Circulation of DENV-23 | 91.51 | 19.87 | - | 421.49 | <0.001 |
| Circulation of DENV-124 | 57.64 | 10.43 | - | 318.60 | <0.001 |
| Circulation of DENV-2 | 43.42 | 9.52 | - | 197.97 | <0.001 |
| Circulation of DENV-1 | 7.59 | 1.41 | - | 40.76 | 0.018 |
| Circulation of DENV-14 | 6.80 | 1.15 | - | 40.37 | 0.035 |
| Circulation of DENV-134 | 2.97 | 0.56 | - | 15.79 | 0.201 |
| Circulation of DENV-13 | 5.84 | 0.57 | - | 59.40 | 0.136 |
| Circulation of DENV-4 | 0.00 | 0.00 | - | 0.00 | 1.000 |
| Circulation of DENV-3 | 0.58 | 0.00 | - | 198.34 | 0.854 |
| Circulation of DENV-34 | 1.49 | 0.01 | - | 154.60 | 0.867 |
| Circulation of DENV-234 | 0.00 | 0.00 | - | 0.00 | 0.999 |
| Circulation of DENV-1234 | 114.16 | 22.90 | - | 568.99 | <0.001 |
| Poverty rate at the regional level | 0.95 | 0.91 | - | 0.99 | 0.013 |
| Total primary health centers over the total population at the regional level | 0.99 | 0.96 | - | 1.03 | 0.797 |

IRRa: Adjusted cumulative incidence ratios per 100,000 population. A mixed negative binomial regression model was used for repeated measures over time.

**S3 Table.** Association between serotype-specific circulation types and cumulative incidence of DS-DWS considering the poverty level variables as a categorical variable of poverty tertiles.

| Variables | Multivariate adjusted analysis |  |  |
| --- | --- | --- | --- |
|  | IRRa | IC 95% | p |
| Circulation of DENV-123 |  |  |  |
| No | Ref. |  |  |
| Yes | 9.38 | 5.13 - 17.14 | <0.001 |
| Circulation of DENV-12 |  |  |  |
| No | Ref. |  |  |
| Yes | 5.90 | 3.53 - 9.86 | <0.001 |
| Circulation of DENV-23 |  |  |  |
| No | Ref. |  |  |
| Yes | 4.36 | 2.28 - 8.35 | <0.001 |
| Circulation of DENV-2 |  |  |  |
| No | Ref. |  |  |
| Yes | 1.94 | 1.01 - 3.73 | 0.047 |
| Circulation of DENV-1 |  |  |  |
| No | Ref. |  |  |
| Yes | 0.27 | 0.10 - 0.76 | 0.013 |
| Circulation of DENV-134 |  |  |  |
| No | Ref. |  |  |
| Yes | 0.11 | 0.04 - 0.28 | <0.001 |
| Poverty tercile at regional level |  |  |  |
| 1 (wealthier) | 14.18 | 4.56 - 44.11 | <0.001 |
| 2 | 8.97 | 4.09 - 19.66 | <0.001 |
| 3 (poorer) | Ref. |  |  |
| Total primary health centers over the total population at the regional level | 1.00 | 0.95 - 1.04 | 0.867 |

IRRa: Adjusted cumulative incidence ratios per 100,000 population. A mixed negative binomial regression model was used for repeated measures over time.

